# A neurocognitive speech taxonomy for voice biomarkers of Alzheimer’s disease

**DOI:** 10.64898/2026.09.23.26363671

**Authors:** Brandon Henley, Zhiyi Yang, Jonathan Sober, Vince Calhoun, Felicia Goldstein, Ihab Hajjar

## Abstract

**INTRODUCTION:** Voice data combined with large language models may detect cognitive impairment, yet an interpretable framework has not been formally established. We developed and validated a scalable and interpretable framework, the Neurocognitive Speech Taxonomy (NST).

**METHODS:** NST maps 314 voice features to 7 neurocognitive domains derived from neuropsychology, speech pathology, neurology and related fields. The domains capture features including articulatory precision, cognitive-linguistic, executive fluency & planning, phonation & laryngeal control, prosodic modulation, lexical-semantic, and morphosyntactic complexity. We evaluated NST structural stability, clinical validity and racial-educational equity in three cohorts (total N = 1,479). Cohort data included demographic and neuropsychological evaluations and AD biomarker status. We tested whether NST scores distinguish cognitive status (unimpaired vs impaired) and AD-biomarker status, track cognitive change, and correlate with AD neuroimaging measures.

**RESULTS:** NST showed stable cross-cohort structure (Mantel *r* = 0.71-0.77) and differentiated MCI (*d* = −0.39) and AD biomarker status (*d* = −0.23). NST tracked longitudinal cognitive change, correlated with hippocampal volume (r up to 0.28, P < .001), and showed smaller racial and educational disparities relative to standard cognitive testing (NST d = 0.03-0.33, MoCA d = 0.37-0.72).

**DISCUSSION:** NST provides a validated neurocognitive construct that facilitates interpretation of AI-driven voice biomarkers of AD. This study was not registered in a public trials registry.

## 1. Introduction

Alzheimer’s disease (AD) and its prodromal stage, mild cognitive impairment (MCI), often go undetected when intervention is most likely to alter the disease course^1,2^. Most neuropsychological tests used in cognitive evaluations vary greatly by educational, cultural, and socioeconomic influences, promoting misclassification in populations with poor quality of education, low acculturation, and impoverished social determinants of health^3–5^. Assessment of spontaneous speech has become an attractive alternative because it is easily obtained, non-obtrusive, brief and allows for repeatability and remote acquisition. Natural language processing and automatic speech recognition of recorded speech have enabled the extraction of linguistic measures reducing reliance on resource-intensive manual transcription and lengthy subsequent discourse analysis.^6,7^ Audio recordings add standardized acoustic analyses to discourse measures expanding cognitive assessment to include multiple dimensions of speech production, including articulation, phonation, speech timing, and prosody. These measures were previously difficult to obtain without advanced computer-aided analyses and have been recently investigated as potential digital biomarkers for cognitive impairment and Alzheimer’s disease.^8–10^ Changes in lexical, semantic, and executive aspects of language are detectable in preclinical stages of AD that are generally undetectable with available commonly used standard cognitive tests. For example, low idea density in early-life writing predicts AD decades later and tracks neurofibrillary tangle burden in the Nun Study^11–13^. However, turning these voice signals into clinically interpretable features that map to specific neurocognitive processes remains a central challenge for their use.

Advances in artificial intelligence, machine learning, computational capacity, and large language models have expanded the ability to extract high-dimensional features and identify subtle patterns in large datasets. When integrated with biological or clinical data, speech-derived measures may improve disease detection, risk stratification, and longitudinal monitoring. However, clinical utility depends on rigorous validation, reproducibility across populations and recording conditions, and demonstrated alignment with established neurocognitive and neuropsychological constructs, which remains a major barrier to interpretable application of AI in cognitive evaluations.^14,15^ To date, much of the connected speech literature has emphasized structural, syntactical, and semantic features that reflect how efficiently information is communicated, spanning the single word level (e.g., use of low vs high frequency words) to the overall richness of the conveyed information (e.g., idea density and cohesiveness). On the other end, newer studies have used LLM embedding which produces a black-box measure difficult to validate and interpret in clinical settings. Speech represents the interplay of numerous processes extending beyond memory, and AD can have heterogeneous presentations apart from memory decline. Although memory impairment is the most common clinical phenotype of AD, patients can present with atypical, non-amnestic syndromes characterized by impairments, for example, in visuospatial processing (i.e., posterior cortical atrophy) and in early-onset autosomal dominant AD cases^16,17^. Neuropsychiatric features may also be the dominant initial presentation as in Mild Behavioral Impairment (MBI)^18^. While neuropsychiatric features have traditionally been linked to frontotemporal dementia or Parkinson’s disease, in vivo evidence also associates them with AD amyloid and tau pathology^19^. MBI may affect the way in which individuals interpret and convey emotional content^20^. Early detection strategies using speech samples should thus assess multiple domains rather than relying exclusively on memory indices. We developed a comprehensive yet interpretable approach to voice analysis addressing the multiple limitations of existing approaches for voice biomarkers in cognition.

Here, we introduce this framework as a two-tier construct that organizes and expands common and novel acoustic, natural language processing (NLP), and large language model (LLM)-derived speech features into the Neurocognitive Speech Taxonomy (NST), which comprises seven neurocognitive domains spanning language-based and motor-speech systems. We validate NST across three independent cohorts (N = 1,479) by evaluating its structural stability, clinical validity, and racial-educational equity. We also compare the effects of race-ethnicity and education on NST performance relative to commonly used cognitive screening tests.

## 2. Methods

### 2.1 Study design and cohorts

Participants in the Brain Stress Hypertension and Aging Research Program (BSHARP; *n* = 491) were recruited in Atlanta^21^. Participants in the Dallas Heart and Mind Study (DHMS; *n* = 711), the most recent collected data from the original Dallas Heart Study, ^22^ and the Post COVID-19 neuro- Cognitive Manifestations and Underlying Mechanisms in Older African Americans (CCARE; *n* = 277) were recruited in the Dallas Fort-Worth multiplex. All participants provided written informed consent under protocols approved by the institutional review boards of Emory University (BSHARP) and UT Southwestern Medical Center (DHMS, CCARE), in accordance with the Declaration of Helsinki. Mild cognitive impairment (MCI) was defined by consensus adjudication based on review of cognitive assessments. AD biomarker positivity (ADpos) was derived from CSF or plasma biomarkers (Table 1). A subset of BSHARP participants (*n* = 402) returned for up to two additional recording visits, yielding 685 longitudinal observations, and 52 CCARE participants returned for one additional visit. For the BSHARP cohort, total hippocampal volume and cortical thickness were also available. Available cognitive tests included Montreal Cognitive Assessment (MoCA) in all 3 studies and Hopkins Verbal Learning Test Revised (HVLT-R) for verbal memory, Trail Making Test for executive function, Boston Naming Test (BNT) for language, and Clinical Dementia Rating (CDR) Scale for overall disease staging in BSHARP cohort.

**Table 1.** Demographic and Clinical Characteristics of Study Cohorts. Values are mean (SD) unless otherwise specified.

|  |  |  |  |
| --- | --- | --- | --- |
| Characteristic | BSHARP<br>(n = 491) | DHMS<br>(n = 711) | CCARE<br>(n = 277) |
| Site | Atlanta | Dallas | Dallas |
| Speech tasks | PD + VF | PD + VF | PD only |
| **NST features** | 314 | 314 | 196 |
| **Age, years** | 65.9 (8.0) | 59.2 (10.8) | 61.7 (7.6) |
| **Education, years** | 15.6 (2.7) | 14.1 (2.6) | 14.8 (2.1) |
| **BMI, kg/m<sup>2</sup>** | 29.1 (7.0) | 32.0 (7.6) | 32.0 (7.1) |
| **MoCA total score** | 23.0 (4.2) | 22.3 (4.4) | 23.9 (3.0) |
| **Male sex, n (%)** | 197 (40.1%) | 415 (58.8%) | 64 (23.1%) |
| **Non-White, n (%)** | 218 (44.4%) | 519 (73.5%) | 253 (100.0%) |
| **APOE4 carrier, n (%)** | 192 (45.6%) | 116 (28.8%) | 98 (37.5%) |
| **MCI, n (%)** | 308 (62.7%) | 196 (28.3%) | 209 (75.5%) |
| **AD biomarker+, n (%)** | 129 (29.7%) | 195 (32.7%) | 40 (15.7%) |

Abbreviations: BSHARP, Brain Stress Hypertension and Aging Research Program; DHMS, Dallas Heart and Mind Study; CCARE, Post COVID-19 neuroCognitive Manifestations and Underlying Mechanisms in Older AfRican AmEricans; PD, picture description; VF, verbal fluency; NST, Neurocognitive Speech Taxonomy; BMI, body mass index; MoCA, Montreal Cognitive Assessment; APOE4, apolipoprotein E $\epsilon$ 4 allele; MCI, mild cognitive impairment; AD, Alzheimer's disease.
AD biomarker+ defined based on CSF, PET, or plasma AD indicators (BSHARP: CSF or PET; DHMS: plasma A $\beta$ 42/40 < 0.161 and p-tau181; CCARE: plasma A $\beta$ 42/40 and p-tau181).
APOE4 carrier percentage calculated among participants with available genotyping.

NST clinical validity was evaluated along two cross-cutting axes applied to the pooled sample. The *“clinical”* axis contrasted participants with MCI against cognitively unimpaired participants. The *biological* axis contrasted ADpos against biomarker-negative participants. Since biomarker status was determined independently of cognitive status, the two axes intersect: biomarker+ participants occurred among those cognitively impaired and unimpaired, and MCI occurred among biomarker+ and biomarker-negative participants. This design allowed NST to distinguish features associated with symptomatic status from those associated with AD pathology.

### 2.2 Voice recording protocol and speech processing

#### Recording protocol

Each participant completed two speech tasks at every visit: (i) a picture description (PD) task reported previously^23^ in which the participant described the “Circus Procession” scene for 120 seconds, and (ii) verbal fluency (VF) tasks comprising letter (phonemic) fluency and category (semantic) fluency. Recordings were obtained in a quiet clinic room using an Apple device and stored for offline analysis. Both PD and VF tasks were collected in BSHARP and DHMS; however, only PD was administered in CCARE.

#### Automated speech processing

Each recording was processed through a pipeline that converts raw audio into participant-only speech (Figure 1). After resampling each audio recording to 16 kHz mono, transcripts were generated by automatic speech recognition (ASR) with word-level time alignment using WhisperX (large-v3). ASR accuracy was benchmarked against human transcription in a subset of 100 BSHARP participants’ baseline recordings, with a median word error rate of 10.79% (range, 1.45%-40.62%). Continuous recordings were segmented into task-specific units, and speaker diarization (pyannote) was then applied to separate the participant from the examiner. All subsequent features were computed from participant-only speech. Transcription, segmentation, and diarization outputs were each passed through an agentic, “LLM-as-judge” quality control (QC) step before passing to the next stage. Specifically, an LLM was used to judge the quality and correctness of output against specific guardrails and rubric for passing QC. The task-specific resampled recordings were loudness-normalized to a fixed root-mean-square reference to mitigate the effects of different recording conditions between the three cohorts (no denoising was applied). Diarization output was passed through an agentic quality-control step that verified and, where needed, corrected the participant-speaker assignments.

**Figure 1.**
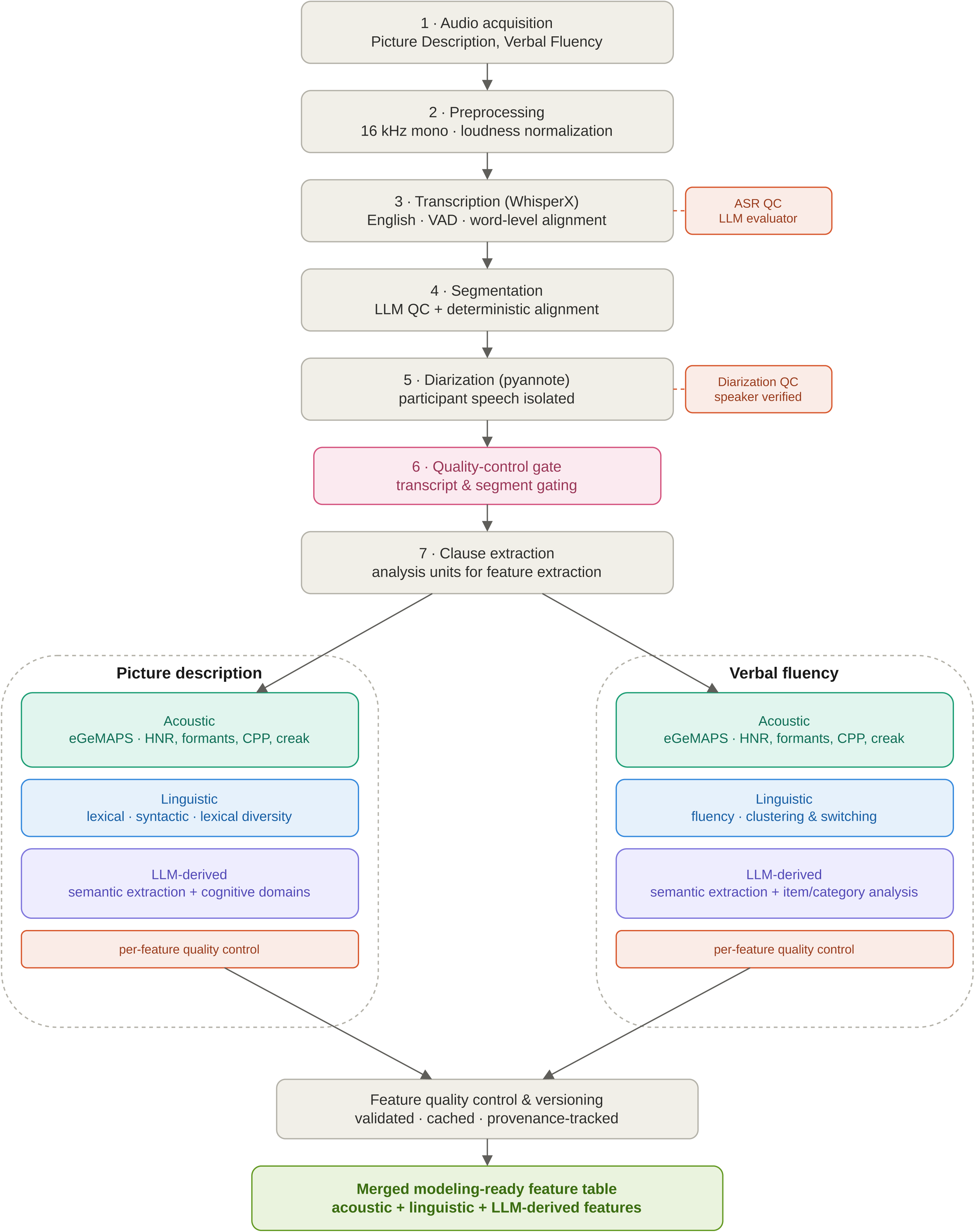
Speech-processing and feature-extraction pipeline. Audio from the picture description (PD) and verbal fluency (VF) tasks is preprocessed (16 kHz mono, loudness-normalized) and transcribed with word-level alignment (WhisperX). The continuous recording is segmented into task units and diarized (pyannote) to isolate participant speech. The transcription, segmentation, and diarization stages are each verified by an LLM quality-control (QC) step (shown in red), and a gating step admits only recordings passing QC. From quality-controlled, participant-only speech, acoustic (eGeMAPS and Praat-based voice-quality and prosodic descriptors, shown in green), linguistic (shown in blue), and LLM-based features (shown in purple) are extracted separately for PD and VF, checked by per-feature QC, and merged into a single feature table.

### 2.3 Feature extraction

From quality-controlled, participant-only speech, we extracted acoustic, linguistic, and LLM-based features from both PD and VF tasks. Our approach combined existing previously described features along with new features that were anchored in neuropsychological and speech production physiology. We describe them below and provide a representative set of features in each category in Table S1.

#### Acoustic features

Acoustic features included the extended Geneva Minimalistic Acoustic Parameter Set (eGeMAPS; 88 parameters spanning frequency, energy, spectral, and temporal descriptors)^8^, extracted with openSMILE^24^. Additionally, we computed acoustic features not obtainable through openSMILE to assess voice-quality and prosodic descriptors with Praat (an open-source phonetics analysis program)^25^, accessed via the Parselmouth Python library^26^: fundamental-frequency and intensity dynamics, spectral tilt, harmonics-to-noise ratio, the first three formants and their dispersion, jitter, shimmer, cepstral peak prominence, and pause statistics. These provided comprehensive coverage of speech production processes supported by both neural and non-neural mechanisms.

#### Linguistic features

Lexical measures were derived using computational linguistic or Natural Language Processing (NLP) and included word and type counts, type-token ratio, Measure of Textual Lexical Diversity (MTLD) and Maas index (lexical diversity), mean word length and syllables per word, content-, function-, and pronoun-word ratios. For the PD task, transcripts were parsed into clauses, from which we derived syntactic-complexity measures (mean dependency depth, subordinate-clause ratio, subject-verb distance, negation and clause counts, mean clause length, and incomplete-clause ratio) and semantic-coherence measures (inter-clause embedding similarity). Fluency and timing measures included speech rate, repetitions, and disfluencies. We also computed semantic-organization features by embedding each PD clause as a sentence-embedding vector and assigning it to the nearest cluster in a pre-trained k-means model, yielding the number of distinct clusters visited, the cluster switch rate, and the mean cluster dwell length per cluster. VF linguistic features included the lexical structure, speech-rate, repetition, and disfluency measures mentioned above.

#### Contextual cognitive-linguistic features

An LLM was used to extract novel features that were non-deterministic or required judgement and context. For instance, it needed to be decided what a word referred to, whether a spoken item is a valid response, or whether a described element reflects a particular cognitive process, none of which can be determined using a fixed algorithm. In this study, we prompted an LLM to read a blinded and deidentified transcript with instructions combined with a structured output template, which was then converted to numeric features using a fixed rule. We passed transcripts for both the PD and VF tasks.

##### Picture description

We created multiple reference points so that the LLM can accurately measure the intended feature. These references were either count-based or attribute-based. For the count-based, we selected 37 canonical scene items that we wanted to detect if the participant mentions them in each transcript (for example, the two elephants, their clothing and actions, the clown, the soldiers, and the title and caption etc.). We instructed LLM to identify which elements of this predefined list were present in the transcript. From this we computed a coverage score (how many distinct canonical items were mentioned) and a repetition score (how many were mentioned more than once). These reflected visual and language processing, visual scanning, attention and potentially ultra-short memory or encoding. We mapped each item to multiple attributes: its appearance, clothing, action (e.g. walking, cycling, laughing etc.), spatial position (in front, behind, above, below etc..), emotion (happy, sad, etc..), and relative size. These were then mapped to nine cognitive constructs (visual perception, visual-spatial processing, perceptual reasoning, attention, memory, executive functioning, abstraction, emotion, and motor perception). Each construct received a score for how many times the participant’s description was mapped to it. The raw scores were then divided by the number of words spoken to obtain a normalized measure for each construct.

##### Verbal fluency

A term was scored valid if the model judged it to meet the task rule: an uttered word or term correctly beginning with the target letter (phonemic) or belonging to the animal or fruit category (semantic), and invalid if it was judged to be an attempted response that did not meet the rule. The LLM was instructed to also flag repetitions of the same term (perseverations), disfluencies (e.g., “um,”, “uh”), and interruption/off-task speech. We computed the number, proportion, and rate of valid responses, and number and proportion of repetitions, disfluencies, and interruptions.

##### Prompt design and reliability

A fixed prompt template was used for each LLM extraction call, into which the transcript was inserted. The LLM was blinded to any additional characteristic of the transcript (such as the source age, sex, race etc.). The output generated was quality checked by comparing the result against predefined guardrails. If the output of the QC call was “pass”, i.e. all guardrails were met, the first LLM call was accepted. Otherwise, a list of errors is provided, which is then passed back and included in the context window of the extraction call to improve the next attempt. Subsequently, deterministic checks were applied to the PD and VF tasks. For PD, it was checked that each listed item belonged to the canonical set. For VF, it was checked that every extracted word was present in the original transcript. Feature generation used greedy decoding (temperature 0.0) to produce deterministic outputs per transcript, cached by a content hash of the prompt and decoding settings. Reliability of the contextual features (canonical-item coverage and repetition) was assessed in a subset of 182 BSHARP PD baseline recordings by using 2 different LLM’s and, measuring between-model agreement using ICC (2,1). Additionally, given LLM potential variability in responses we calculated within-model reproducibility by running five repeated features using the two LLMs and calculating intra-model agreement. These results are shown in Table S2 All features were assembled into a single table prepared for modeling. Extraction code and prompts are available in a versioned private repository upon request.

### 2.4 NST taxonomy description

We organized the features into a two-tier neurocognitive speech taxonomy where each feature was mapped to one of seven candidate neurocognitive domains (Tier 1) derived from neuropsychology, speech pathology, neurology and related fields and covered voice production characteristics, neural and motor controls, as well as possible cognitive processes and representations derived from the transcripts. Features were also mapped to a second-tier domain with more granular descriptors within each domain reflecting a more specific process. The seven domains are shown in (Figure 1):

1. *Articulatory Precision* captures how accurately and consistently the vocal tract shapes speech sounds and resonance control through formant structure and spectral shape.
2. *Cognitive-Linguistic* captures the informational content of the transcript reflecting what the speaker notices, retrieves, organizes, and communicates rather than how sound is produced.
3. *Executive Fluency & Planning* captures the efficiency and temporal organization of speech production and reflects indexing lexical search, pausing, timing regularity, repetitions, and the ability to maintain a continuous verbal stream that is not interdigitated with a lot of pauses.
4. *Phonation & Laryngeal Control* captures how the larynx and vocal folds generate and regulate sound, reflecting pitch control and stability, harmonic quality, jitter and shimmer, and overall.
5. *Prosodic Modulation* captures how the speaker modulates vocal energy focusing on loudness-related prosody such as intensity level, range, dynamics, and temporal density of energy peaks of the sound wave.
6. *Lexical-Semantic* Domain captures the richness, breadth, and sophistication of words used, reflecting lexical diversity, vocabulary size, and the quality of the word-level repertoire.
7. *Morphosyntactic Complexity* captures the complexity and organization of sentences, reflecting clause structure, syntactic span, sentence completeness, and the content organization.

### 2.5 Domain score computation

NST domain scores for tiers 1 and 2 were computed using Principal Component Analysis (PCA) of the domain-specific features after being standardized, and the first principal component was extracted. We preferred PCA(1) to a simple mean of z-scored features because NST domains included features with heterogeneous variance and pairwise correlation structure; PCA yields a data-driven composite that weights features by shared covariance and reduces redundancy especially with collinear indicators.^27^ In addition to the individual domains, a global NST composite score was computed as the sum of the seven tier-1 domain scores, providing a single summary measure.

### 2.6 Structural validity and clinical utility

We evaluated the internal coherence of NST by computing the feature-to-feature correlation matrix and calculating the within-domain vs between-domain absolute correlation ratios (coherence).^28^ Features were also projected into two dimensions using multidimensional scaling (MDS) applied to the distance matrix to assess the degree of NST domain separation.^29^ Cross-cohort stability was evaluated with Mantel tests using 1,000 permutations to compare condensed distance matrices across all cohort pairs.^30^ This evaluated whether the overall pattern of relationships among features within each domain was similar across different cohorts. Clinical utility was conducted by quantifying group differences with Cohen’s *d* of NST tier 1 and tier 2 domain scores between MCI and AD biomarker positive groups. Results were pooled across cohorts using fixed-effect inverse-variance meta-analysis. Benjamini-Hochberg false discovery rate (FDR) correction was applied at q < 0.05 to adjust for the multiple testing. In the longitudinal data, NST change was assessed using linear mixed-effects models (LMEM) trajectories that were stratified by MCI and ADpos. Racial and educational effects were assessed by the effect size (Cohen’s *d*) between White vs Black/African American mean NST domain scores and Pearson correlations with years of educational attainment vs. MoCA scores.

## 3. Results

### 3.1 Sample Description

Participant characteristics are summarized in Table 1. BSHARP sample (*n* = 491) had a mean age of 65.9 (8.0) years, was 40% male, 56% White, and had a mean MoCA of 23.0 (4.2); 63% met criteria for MCI. DHMS sample (*n* = 711) had a mean age of 59.2(10.8), 59% were male, 27% White and 28% met criteria for MCI. CCARE (*n* = 277) had a mean age of 61.7 (7.6), was entirely African American, predominantly female (77%), with 76% meeting MCI criteria. CCARE included only picture-description recordings. The NST taxonomy assigned all 314 features to 7 tier-1 domains and 50 tier-2 categories (Figure 2).

**Figure 2.**
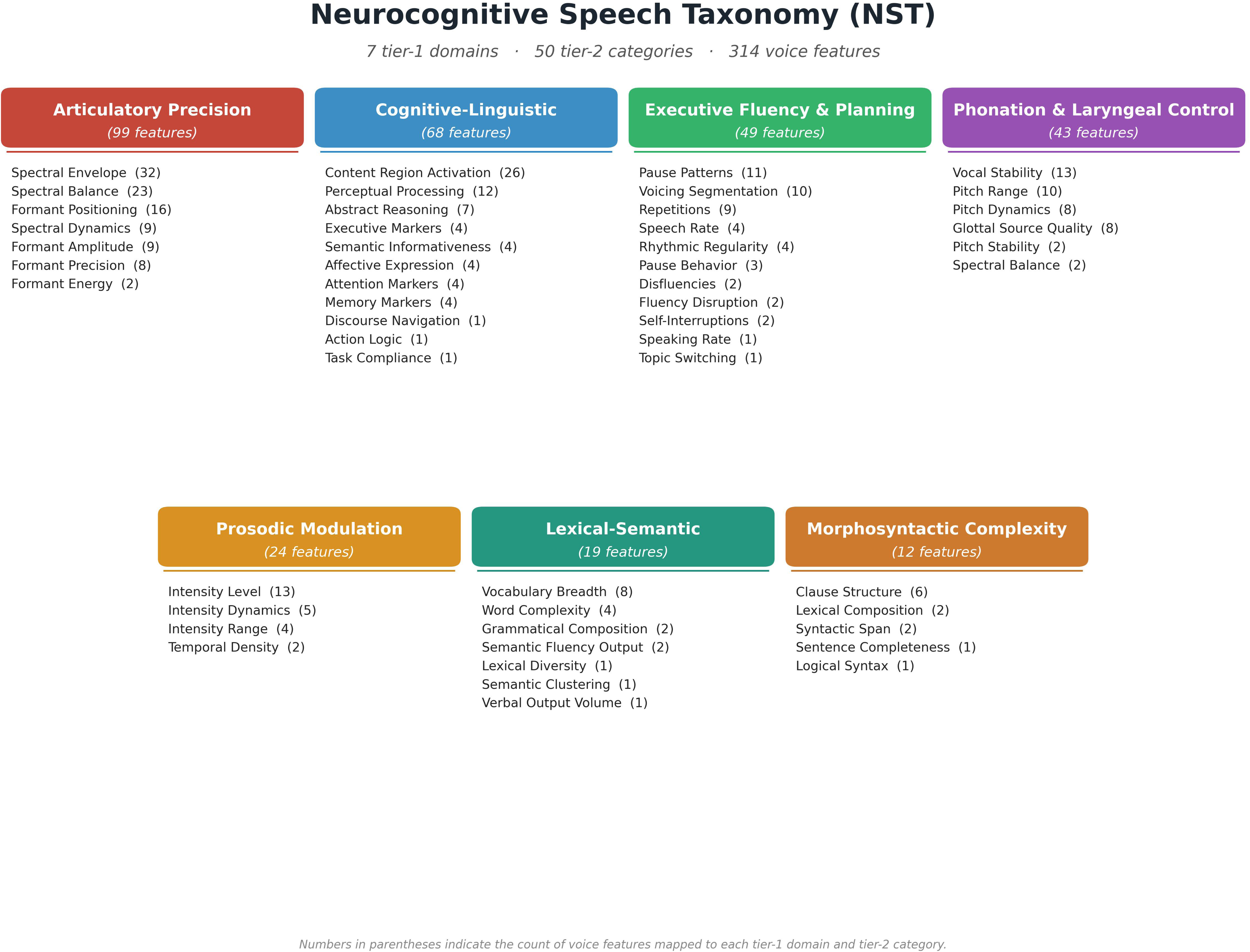
The 314 voice features are organized into seven tier-1 neurocognitive domains and 50 tier-2 categories. The total feature count in each domain and each category shown in parentheses.

### 3.2 Data-Driven Construct Validity

The NST domain data structure was supported by three complementary analyses pooling all three cohorts (N = 1,479; Figure S1). Feature-by-feature Pearson correlation matrices showed block-diagonal structure along the NST domain ordering (Figure S1, panel A). Within-domain mean correlations exceeded between-domain mean correlations across all seven domains (Figure S1, panel B; Table S3), with Phonation & Laryngeal Control showing the highest coherence and Articulatory Precision the lowest. Multidimensional scaling showed visible domain grouping in twodimensional space (Figure S1, panel C), with language-based domains (Cognitive- Linguistic, Lexical-Semantic, Morphosyntactic) and motor-speech domains (Articulatory Precision, Phonation) forming 2 separate regions. This domain structure is replicated across all three cohorts with the Mantel test which measures the similarity of the domain structures between pairs of cohorts ranging between 0.71-0.77 (all *p* < 0.001; Table S4) even when one cohort had only one task (PD).

### 3.3 Clinical Validity

Clinical validity was assessed by comparing NST scores along 2 dimensions (MCI vs cognitively unimpaired and AD positive vs negative groups). Meta-analytic effect sizes from the combined sample are presented in Table 2 and Figure 3. NST composite score was significantly different by MCI (*d* = −0.39, *p* < 0.001) and AD biomarker positivity (*d* = −0.23, *p* < 0.001), both surviving FDR correction. Lexical-Semantic (*d* = −0.41, *p* < 0.001) and Executive Fluency & Planning (*d* = −0.41, *p* < 0.001) showed the largest MCI effects. Lexical-Semantic and Cognitive-Linguistic domains were significantly different by AD biomarker positivity (p < 0.001). Per-cohort domain-level effect sizes are provided in Table S5 (tier 1) and Table S6 (tier 2). NST composite and Lexical Semantic, executive fluency but not Motor Speech domains showed significant MoCA correlations in BSHARP and DHMS (Figure 4). In CCARE (PD-only), correlations were attenuated but directionally consistent, with Lexical-Semantic (*r* = 0.16, *p* = 0.007) reaching significance.

**Figure 3.**
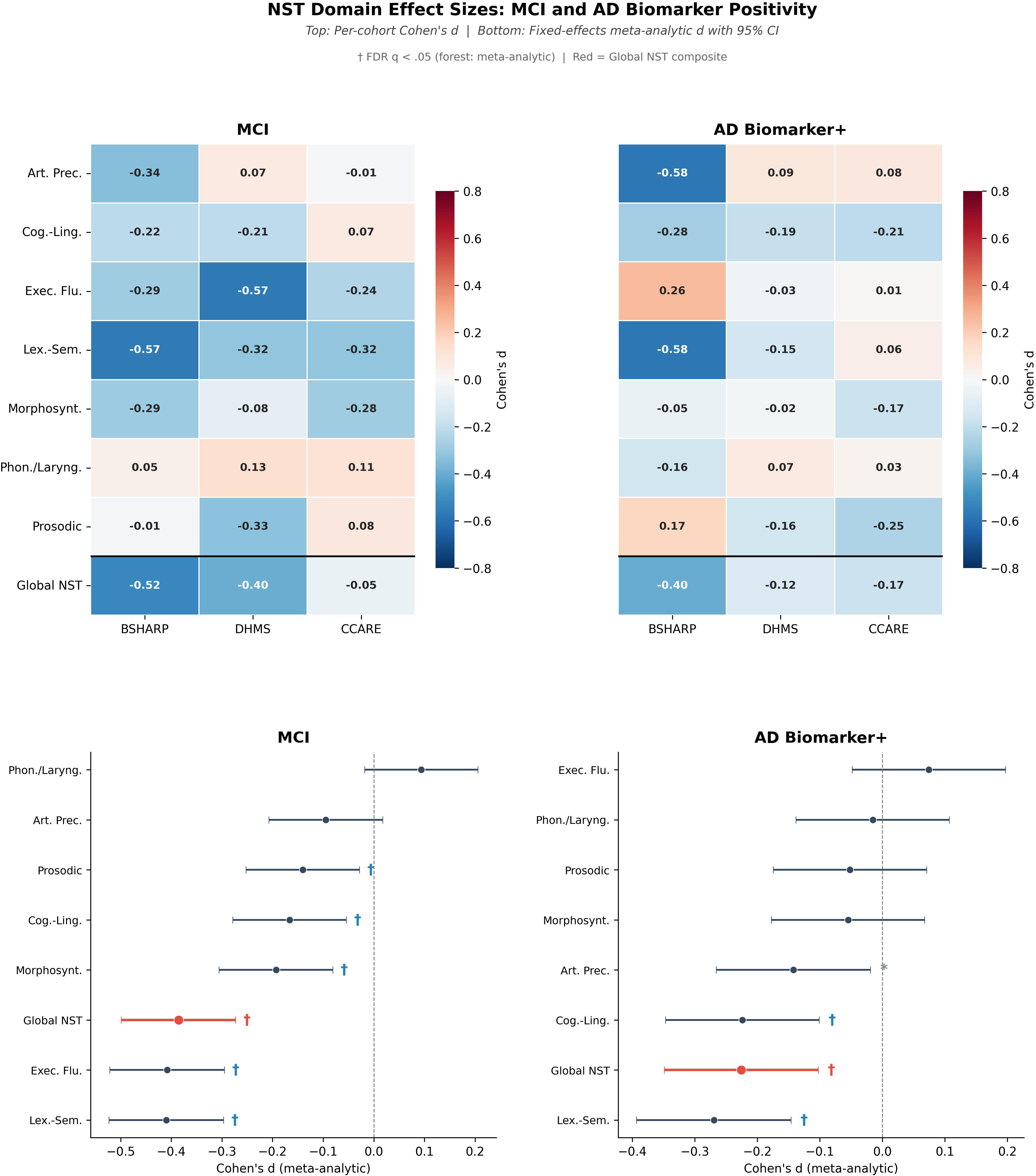
*Top*: Cohen’s d for each tier-1 domain and the global NST composite in the BSHARP, DHMS, and CCARE for MCI (left) and AD-biomarker positive (right) contrasts. Negative values mean lower scores in the impaired/biomarker+ group. *Bottom:* fixed-effects meta-analytic d with 95% confidence intervals in order of effect size. Global NST composite shown in red. †FDR q < 0.05.

**Figure 4.**
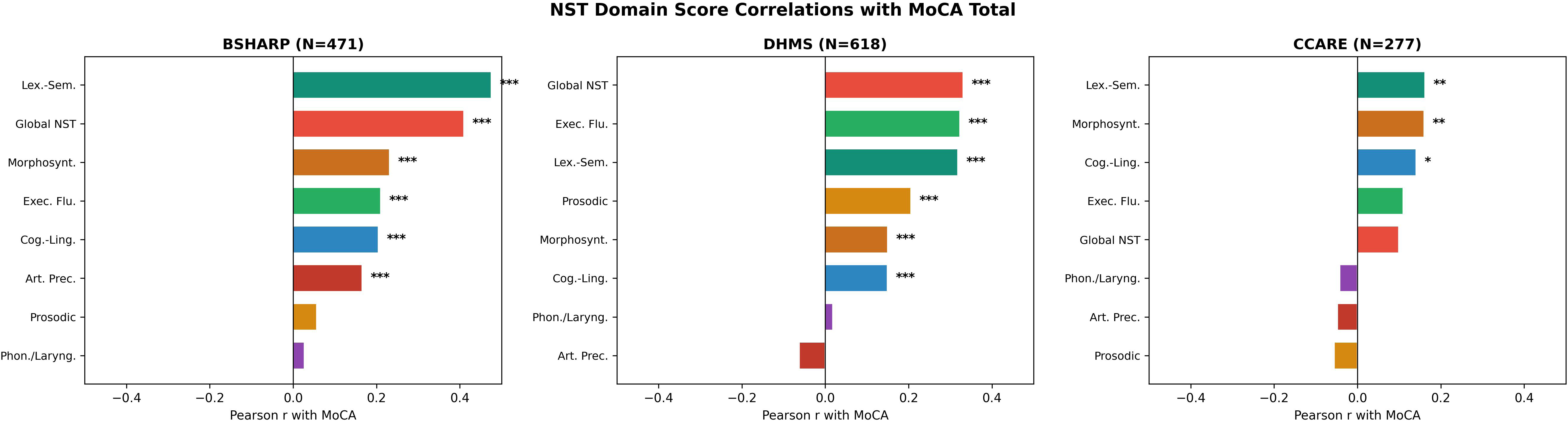
Pearson correlations between each tier-1 domain (and the global NST composite) and MoCA Total in BSHARP, DHMS, and CCARE. * p < 0.05, ** p < 0.01, *** p < 0.001.

**Table 2.** Fixed-effects meta-analysis pooling Cohen's d across BSHARP, DHMS, and CCARE cohorts. Negative d indicates lower scores in the impaired group.

| Domain | MCI |  |  |  | AD Biomarker+ |  |  |  |
| --- | --- | --- | --- | --- | --- | --- | --- | --- |
|  | d (SE) | I<sup>2</sup> | p | FDR q | d (SE) | I<sup>2</sup> | p | FDR q |
| **Articulatory Precision** | -0.10<br>(0.057) | 82% | 0.098 | 0.0297 | -0.14<br>(0.063) | 92% | 0.024 | 0.0234 |
| **Cognitive-Linguistic** | -0.17<br>(0.057) | 41% | 0.004 | 0.0203 * | -0.22<br>(0.063) | 0% | < 0.001 | 0.0141 * |
| **Executive Fluency and Planning** | -0.41<br>(0.058) | 70% | < 0.001 | 0.0031 * | 0.07<br>(0.063) | 57% | 0.234 | 0.0344 |
| **Lexical-Semantic** | -0.41<br>(0.058) | 55% | < 0.001 | 0.0016 * | -0.27<br>(0.063) | 86% | < 0.001 | 0.0063 * |
| **Morphosyntactic Complexity** | -0.19<br>(0.057) | 40% | < 0.001 | 0.0172 * | -0.06<br>(0.063) | 0% | 0.383 | 0.0391 |
| **Phonation and Laryngeal Control** | 0.09<br>(0.057) | 0% | 0.100 | 0.0312 | -0.01<br>(0.063) | 29% | 0.808 | 0.0469 |
| **Prosodic Modulation** | -0.14<br>(0.057) | 78% | 0.014 | 0.0219 * | -0.05<br>(0.063) | 72% | 0.410 | 0.0422 |
| **Global NST** | -0.39<br>(0.058) | 74% | < 0.001 | 0.0047 * | -0.23<br>(0.063) | 55% | < 0.001 | 0.0125 * |

FDR q < 0.05.
Abbreviations: d, Cohen's d (standardized mean difference); SE, standard error; I<sup>2</sup>, heterogeneity statistic (percentage of variance due to between-study differences); FDR, false discovery rate; MCI, mild cognitive impairment; AD, Alzheimer's disease; NST, Neurocognitive Speech Taxonomy.
Global NST is the sum of all 7 domain scores.

### 3.4 Longitudinal Change

Linear mixed-effects models within the full study sample showed progressive decline in all NST domains except for Prosodic Modulation and Executive Fluency planning which showed improvements. (Figure S2, Panel A). However, when separated by MCI and ADpos groups, those with preclinical AD demonstrated the greatest annual decline in the NST composite, Articulatory Precision and Cognitive Linguistic domains (Figure S2, Panel B).

### 3.5 Impact of Race and Education on NST

All seven NST domain scores showed smaller absolute disparities between the 2 racial groups than MoCA in both cohorts. Cognitive-Linguistic (*d* = 0.03) and Lexical-Semantic (*d* = 0.13) showed the smallest racial differences in BSHARP. In DHMS, where the MoCA gap was large, all seven NST domains remained below *d* = 0.33. Executive Fluency showed the largest domain disparity (*d* = 0.30-0.33), but still below MoCA. Education correlated moderately with MoCA in both BSHARP and DHMS but showed weaker and more domain-specific associations with NST scores (Table S7). Articulatory Precision, Phonation, and Prosodic Modulation were largely education independent.

### 3.6 Correlation between NST Scores and traditional cognitive tests and Neuroimaging Measurements

NST domain scores were compared with five neuropsychological measures (MoCA Total, HVLT delayed recall, BNT, Trails B-A, and CDR Total; Figure 5). All correlations were in the expected direction: positive with MoCA, HVLT delayed recall, and BNT, and negative with Trails B-A and CDR. The Lexical-Semantic domain showed the strongest associations (MoCA r = 0.49, HVLT delayed recall r = 0.44, BNT r = 0.35, Trails B-A r = −0.34, CDR r = −0.29, all p < 0.001). Cognitive-Linguistic, Executive Fluency & Planning, Morphosyntactic Complexity, and Articulatory Precision all correlated with MoCA (r = 0.16-0.23, p < 0.001). Phonation & Laryngeal Control and Prosodic Modulation were weakly correlated with cognitive tests. In the subset of participants with available structural MRI (n = 386, BSHARP), NST domain scores showed weak-to-moderate positive associations with regional brain structure (Figure 5). In particular, hippocampal volume was significantly associated with five domains: Lexical-Semantic (r = 0.28, p < 0.001), Cognitive-Linguistic (r = 0.17, p = 0.001), Phonation & Laryngeal Control (r = 0.13, p = 0.011), Articulatory Precision (r = 0.12, p = 0.022), and Executive Fluency & Planning (r = 0.12, p = 0.017). Cortical thickness was significantly associated with three domains: Executive Fluency & Planning (r = 0.21, p < 0.001), Prosodic Modulation (r = 0.15, p = 0.003), and Lexical-Semantic (r = 0.10, p = 0.043). All significant correlations were positive, indicating higher domain scores correspond to greater hippocampal volume and cortical thickness.

**Figure 5.**
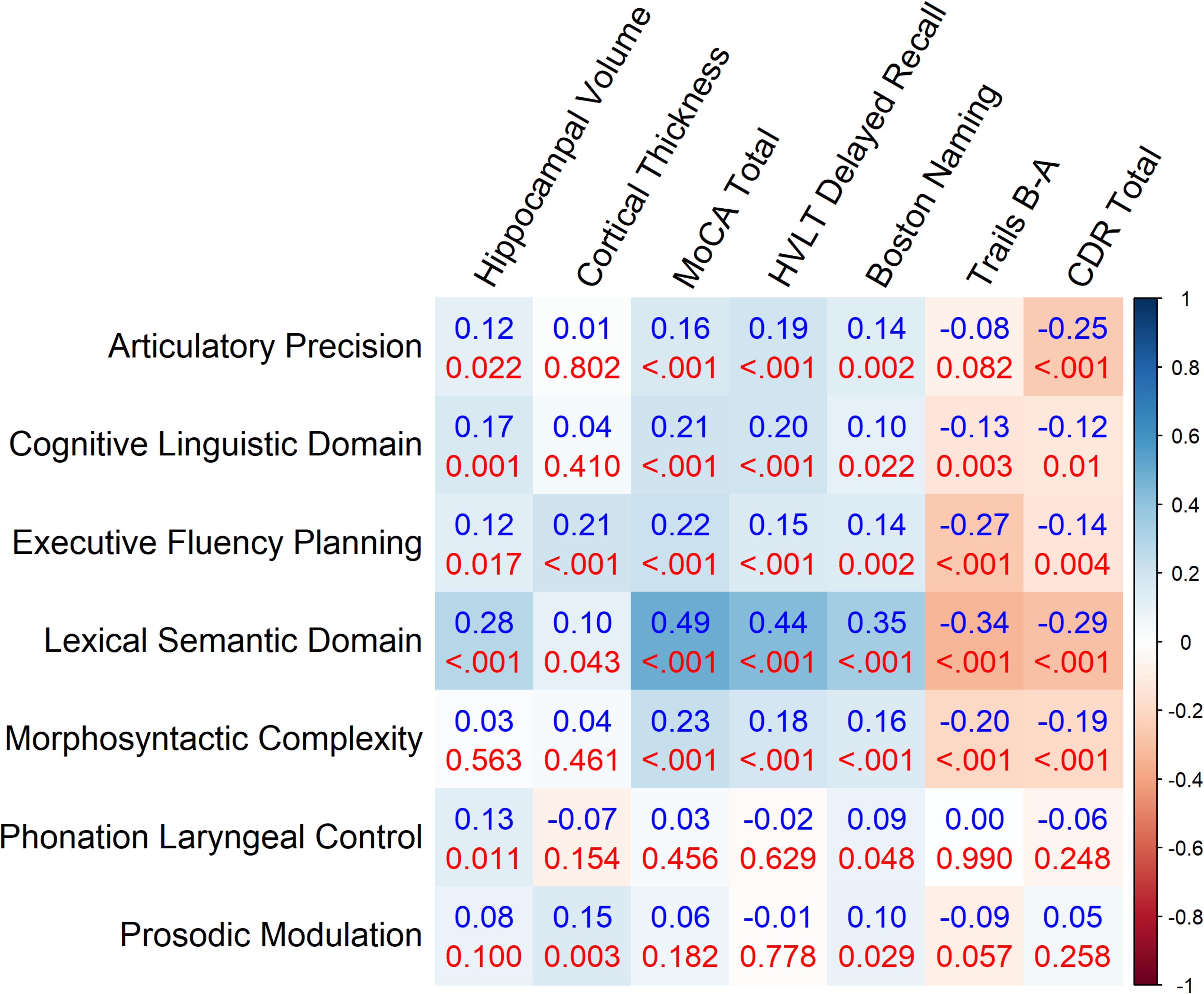
Pearson correlations between each tier-1 domain and MoCA Total, HVLT delayed recall, BNT, Trails BA, CDR Total, hippocampal volume, and cortical thickness. In each cell, the top value is the correlation coefficient, and the bottom value is the p-value.

## Discussion

We developed and validated the Neurocognitive Speech Taxonomy, a framework that organizes extracted features from a brief audio recording into seven neurocognitive domains to make these digital biomarkers more interpretable. NST is intended as an interpretive layer that can be used in predictive models by identifying which speech-cognitive systems contribute most to either cognitive impairment or AD pathological prediction. The domain structure was stable across cohorts and showed smaller demographic disparity relative to MoCA.

Executive and lexical domains were more associated with symptomatic status (MCI), whereas semantic, content-based domains were more associated with AD-biomarker status^31^. We interpret these modest contrasts as a pattern warranting further study. Executive Fluency & Planning and Lexical-Semantic illustrate this distinction as the domains most sensitive to symptomatic status. Executive Fluency separated MCI from normal cognition, but not biomarker+ from biomarker-negative participants, suggesting that reduced executive fluency indexes the symptomatic (MCI) stage rather than the underlying AD pathology. This observation is consistent with evidence that the switching component of verbal fluency is frontally and executively mediated whereas clustering reflects semantic memory,^32^ and that semantic fluency declines more than phonemic fluency in AD,^33^ indexing both executive and semantic processes. Unlike Executive Fluency, the Lexical-Semantic domain showed the largest effect on both axes, indicating that this domain exhibits a dual loading, tracking both the symptom and pathology axes. In general, the NST domains were separable but not orthogonal: within-domain correlations exceeded between-domain correlations (Figure S1B), but between-domain correlations were non-zero, consistent with shared processes across domains. For example, lexical fluency draws on both executive and semantic control. Therefore, domain-level effects should be interpreted as partially overlapping rather than statistically independent.

Along the pathology axis, the domains associated with AD-biomarker status were Lexical-Semantic and Cognitive-Linguistic. Because the Lexical-Semantic domain combines measures of both lexical diversity and semantic content, its association with biomarker status could in principle reflect either one. A prior study, however, linked amyloid to fewer specific and content words in cognitively unimpaired adults but not type-token ratio, a measure of vocabulary diversity.^34^ Another study reported reduced content words and category fluency but no change in diversity measures. ^35^ In general, these observations are consistent with the well-established decline in semantic memory in AD,^36^ semantic deficits as one of the earliest detectable language changes,^37^ and the association of connected-speech and semantic memory in AD.^38,39^ A single global score would combine the domains that relate more to symptomatic impairment with those that are associated more with pathology, obscuring the distinction that makes the taxonomy clinically informative.

The association between NST domains and structural imaging was modest but reinforced the construct validity of the taxonomy. Hippocampal volume is an early marker of Alzheimer’s neurodegeneration and was most strongly related to the Lexical-Semantic domain, consistent with the medial temporal contribution to semantic and lexical retrieval. Lexical-Semantic and Executive Fluency & Planning domains showed broad structural associations, suggesting that these domains capture AD-relevant neurodegeneration. Since these correlations were weak and cross-sectional, they should be interpreted as supporting construct validity rather than establishing a clear relationship between structure and cognitive function.

The NST domains appear to serve different clinical roles. The motor-speech domains (Articulatory Precision, Phonation) were not sensitive cross-sectionally, and Articulatory Precision showed the steepest longitudinal decline. This is consistent with the traditional observation that articulation and phonological processing are relatively preserved in early AD^40,41^, although recent work reports earlier motor-speech changes^42^, and with acoustic features predicting incident dementia in asymptomatic adults.^43^ By contrast, Lexical-Semantic and Executive Fluency were sensitive cross-sectionally and correlated with MoCA. Therefore, these patterns suggest distinct clinical roles: cross-sectional screening, longitudinal progression monitoring.

NST domains showed smaller demographic disparities than MoCA. In the BSHARP and DHMS cohorts (CCARE only had one racial group), all seven domains exhibited smaller absolute racial differences than MoCA, with Cognitive-Linguistic and Lexical-Semantic the smallest. Education correlated with MoCA but only weakly and selectively with NST domains, and was essentially independent of Articulatory Precision, Phonation, and Prosodic Modulation. This observation is notable because traditional cognitive tests show race- and education-related performance gaps,^44^ AD prediction models can propagate racial disparities,^45^ and speech-based detectors are themselves demographically biased.^46,47^ A smaller disparity is clinically meaningful only if it does not simply reflect reduced sensitivity. Two observations indicate that it does not: the domains with the smallest demographic gaps (Lexical-Semantic, Cognitive-Linguistic) were also among those with the largest and most reproducible clinical effects, and speech-based screening keeps better operating characteristics than brief cognitive tests.^48^ Therefore, we interpret reduced disparities as fairer measurement rather than insensitivity, though this remains a measurement property that requires future confirmation.

The findings and interpretations above have several translational implications. Because the proposed domains separate a symptomatic axis from a pathology axis, an NST *profile*, rather than a single score, may help distinguish cognitive symptoms from underlying AD biology. Such profiling is consistent with prior work that combines speech with fluid or imaging biomarkers.^49,50^ The NST framework is also low-burden and scalable for deployment, since speech can be collected remotely and reproducibly.^51,52^ As we have seen, different domains may serve different roles. Separately, by mapping heterogeneous features onto cognitively meaningful domains, NST offers a shared vocabulary that may improve cross-study comparability and reduce reliance on black-box outputs.^53^ Given its smaller demographic disparities, NST may be particularly valuable in primary-care settings where misclassification by traditional cognitive screening is driven by other factors. We propose these as candidate applications requiring prospective validation before clinical use.

Several important limitations qualify our findings. First, LLM-derived features were highly reproducible within the same LLM (median ICC 0.92), but repetition was moderately reproducible across models (ICC 0.32). Second, the cross-sectional effect sizes were modest, so NST is best positioned as a tool for interpretation complementary to, not a surrogate for, diagnostic assessment. Modest preclinical effects are not uncommon for speech biomarkers.^48,51,54^ Third, several domains showed large between-cohort heterogeneity (I^2^ up to 92%), so pooled effect sizes should be considered alongside their I^2^. For this reason, fixed-effects pooling estimates should be interpreted cautiously for highly heterogeneous domains, and random-effects pooling may be preferable. The Lexical-Semantic domain combines lexical-diversity and semantic-content measures, which can dissociate etiologically. Since our composite did not separate these contributions, its association should be understood as reflecting either component rather than a specific one. CCARE contributed picture description only, so that 196 of the 314 features were available for this cohort. This meant under-measurement of the domains dependent on motor-speech and verbal-fluency, limiting their cross-cohort comparability. All cohorts were English-speaking and US-based; although the domain structure and lexical-semantic findings have cross-linguistic support, generalization of NST itself to other languages and recording conditions remains to be tested. Finally, inter-participant differences in speech due to accent, dialect, and prosody may influence domain scores, particularly the motor-speech domains; this study did not consider these effects on NST and warrants dedicated study.

We conclude that NST reframes voice biomarkers of AD toward a more interpretable measurement tool, organizing 314 speech features into a condensed set of seven clinically relevant neurocognitive domains with a stable cross-cohort structure, differential sensitivity to cognitive symptoms and AD pathology, and smaller demographic disparities than standard cognitive screening. Pending prospective validation, NST offers a scalable, interpretable foundation for speech-based cognitive assessment.

## Acknowledgments

The authors thank the participants and staff of the BSHARP, DHMS, and CCARE studies.

## Consent Statement

All participants provided written informed consent in accordance with the Declaration of Helsinki. Study protocols were approved by the institutional review boards of Emory University (BSHARP) and UT Southwestern Medical Center (DHMS, CCARE).

## Data Availability Statement

Data and code used in this manuscript are available upon request.

