## Supplementary Material for "A neurocognitive speech taxonomy for voice biomarkers of Alzheimer’s disease"

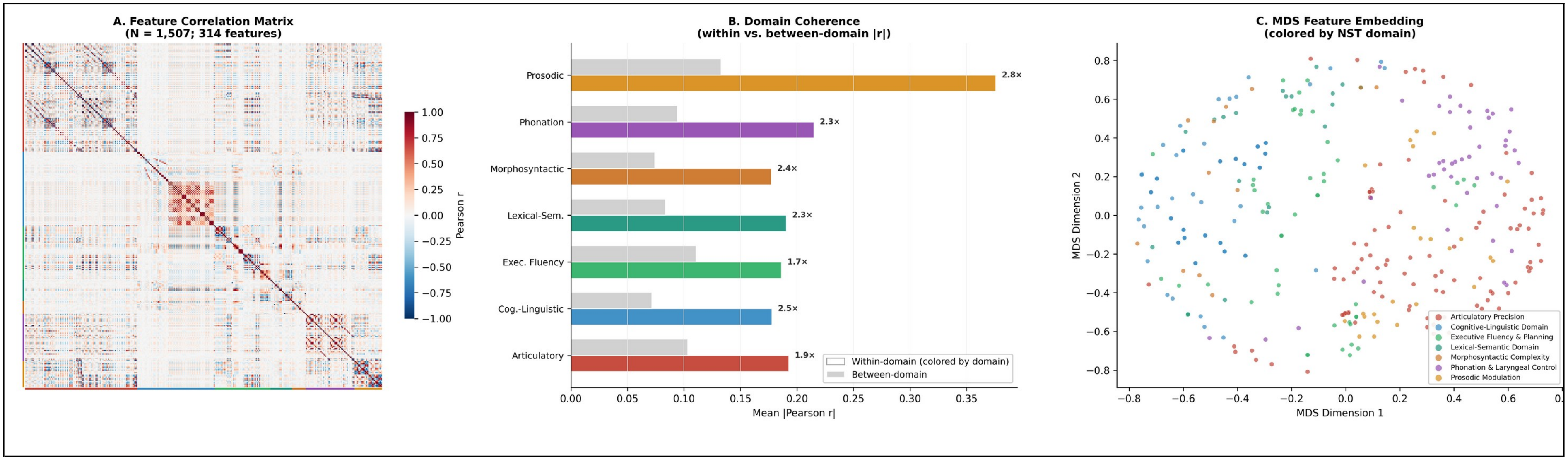


Figure S1. Structural validation of the NST taxonomy pooled across the 3 cohorts.
(A) Feature correlation matrix ordered by NST tier-1 domain. (B) Within-domain vs between-domain mean |Spearman r| per domain. (C) Two-dimensional MDS embedding of the 1 − |r| feature distance matrix, colored by NST tier-1 domain.

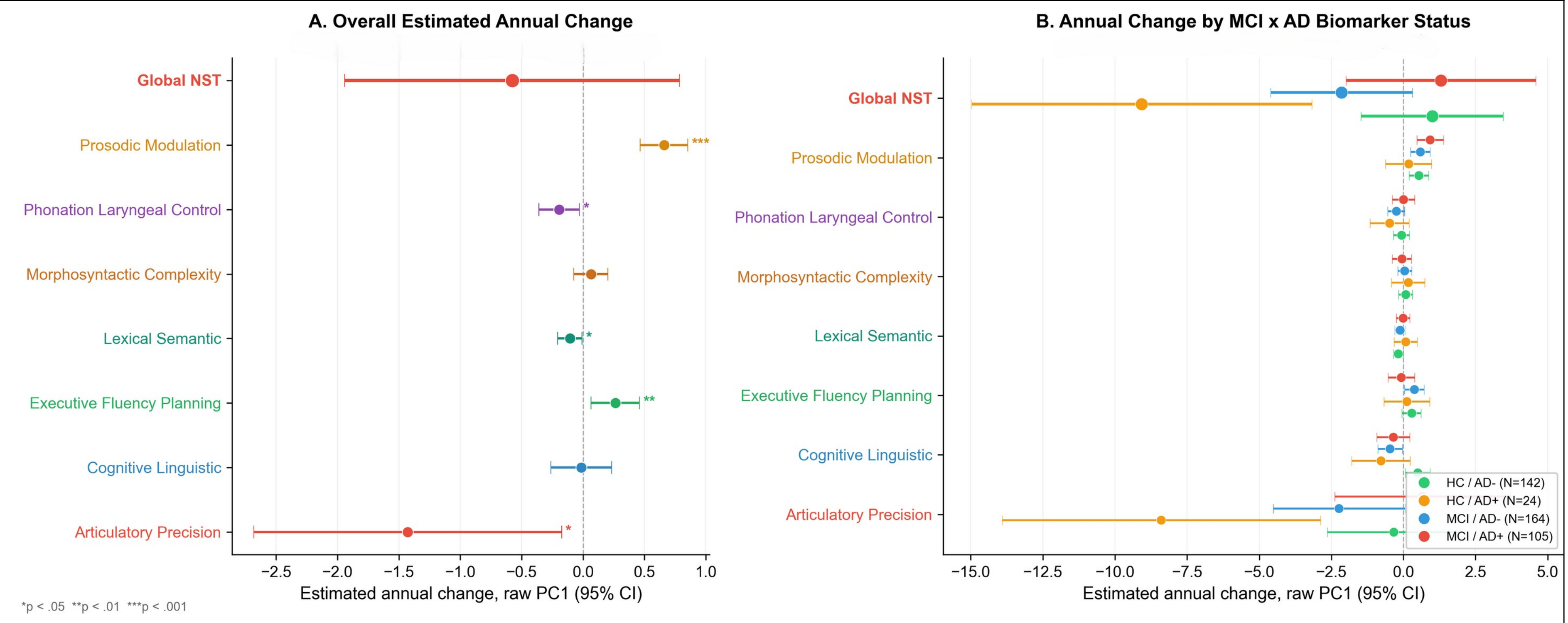


Figure S2. Longitudinal NST domain score changes in BSHARP (N = 402, visits 1–3) using raw PCA direction without sign alignment.

(A) Overall estimated annual change per domain from a linear mixed model). (B) Annual change stratified by MCI × AD biomarker status. Points show estimated slopes with 95% confidence intervals; †p < 0.1, *p < 0.05, **p < 0.01.

Table S1. Representative NST features and their definitions. For each category (acoustic, linguistic, and LLM-derived)

| category | feature | definition |
|---|---|---|
| Acoustic (custom) | Harmonics-to-noise ratio | Mean harmonics-to-noise ratio in dB. |
| Acoustic (custom) | Jitter | Local jitter expressed as a percentage of F0. |
| Acoustic (custom) | Shimmer | Local shimmer expressed as a percentage of amplitude. |
| Acoustic (custom) | First formant (F1) | Mean frequency of first formant (F1) in Hz. |
| Linguistic (PD and VF) | Lexical diversity (MTLD) | Measure of textual lexical diversity (MTLD), computed across the picture-description or verbal-fluency transcript. |
| Linguistic (PD and VF) | Speech rate | Words spoken per second. |
| Linguistic (PD) | Syntactic dependency depth | Average depth of syntactic dependency trees (syntactic complexity). |
| Linguistic (PD) | Incomplete clause ratio | Proportion of clauses that are incomplete or fragmentary. |
| Linguistic (PD) | Inter-clause semantic similarity | Mean pairwise cosine similarity between clause embeddings in the picture-description transcript (semantic coherence across the narrative). |
| Linguistic (VF) | Repetition rate | Rate of repeated items relative to total words. |
| Linguistic (VF) | Disfluency rate | Rate of filled pauses and related disfluencies relative to words in the verbal-fluency transcript. |
| Linguistic (VF) | Mean repeat distance | Average distance in words between repeated items. |
| PD LLM | Item coverage | Proportion of canonical circus-picture content items mentioned at least once in the LLM-extracted item list. |
| PD LLM | Item repetitions | Proportion of redundant item mentions in the LLM-extracted picture-description sequence. |
| PD LLM | Executive functioning | Proportion of possible executive-functioning credits earned from LLM-scored picture attributes (relative to the domain maximum). |
| VF LLM | Valid response rate | Proportion of spoken words that are valid category responses in the verbal-fluency task, as identified by the LLM. |
| VF LLM | Perseverations | Proportion of verbal-fluency responses that are perseverative repetitions, as |

| | | |
|---|---|---|
| | | identified by the LLM. |
| VF LLM | Disfluencies | Proportion of verbal-fluency responses that are disfluencies (e.g., fillers or false starts), as identified by the LLM. |

Table S2. Reliability of LLM-Derived Picture Description Features.

| Construct | Within LLM 1 | Within LLM 2 | Between LLM 1 & 2 |
|---|---|---|---|
| Item coverage | 0.97 | 0.96 | 0.74 |
| Repetitions | 0.86 | 0.82 | 0.32 |

Values are intraclass correlation coefficients (ICC(2,1)) in a subset of 182 BSHARP baseline recordings, for within-model reproducibility across five repeated extractions and between model agreement. Item coverage was reproducible with and across models, whereas repetition counts were reproducible within models but weakly consistent across models.

Table S3. Within-Domain Feature Coherence Across Cohorts. Values shown as within-domain mean |r|/between-domain mean |r| (ratio).

| **Cohort** | **Articulatory Precision** | **Cognitive-Linguistic** | **Executive Fluency** | **Lexical-Semantic** | **Morpho-syntactic** | **Phonation/ Laryngeal** | **Prosodic Modulation** |
|---|---|---|---|---|---|---|---|
| **BSHARP** | 0.16/0.10 (1.7) | 0.18/0.07 (2.5) | 0.24/0.11 (2.2) | 0.20/0.09 (2.2) | 0.20/0.08 (2.6) | 0.23/0.10 (2.4) | 0.34/0.11 (3.1) |
| **DHMS** | 0.19/0.10 (2.0) | 0.18/0.07 (2.6) | 0.25/0.11 (2.2) | 0.19/0.08 (2.4) | 0.18/0.08 (2.3) | 0.24/0.10 (2.5) | 0.34/0.12 (2.9) |
| **CCARE** | 0.21/0.11 (1.9) | 0.20/0.10 (1.9) | 0.24/0.15 (1.6) | 0.23/0.14 (1.6) | 0.21/0.10 (2.1) | 0.27/0.13 (2.0) | 0.64/0.15 (4.3) |

Ratio > 1 indicates features within a domain correlate more strongly with each other than with features in other domains, supporting structural coherence.

Abbreviations: r, Pearson correlation coefficient; BSHARP, Brain Stress Hypertension and Aging Research Program; DHMS, Dallas Heart and Mind Study; CCARE, Post COVID-19 neurocognitive Manifestations and Underlying Mechanisms in Older African Americans.

Table S4. Cross-Cohort Structural Stability: Mantel Test of Feature Correlation Matrices. Mantel test assesses whether feature-by-feature correlation matrices are structurally similar across cohorts.

| Cohort Pair | n Features | Mantel r | p |
|---|---|---|---|
| **BSHARP-DHMS** | 313 | 0.7541 | 0.001 |
| **BSHARP-CCARE** | 195 | 0.7086 | 0.001 |
| **DHMS-CCARE** | 195 | 0.7705 | 0.001 |

High r values indicate that the correlation structure among NST features is preserved across independent samples.

p-values computed via 9,999 permutations.

BSHARP–CCARE and DHMS–CCARE comparisons use the 195 PD-only features available in CCARE.

Abbreviations: r, Mantel correlation coefficient; NST, Neurocognitive Speech Taxonomy; PD, picture description.

Table S5. Per-Cohort Effect Sizes for NST Domain Scores by Clinical Outcome (Tier 1). Values are Cohen's d (standardized mean difference) comparing outcome-positive vs outcome-negative groups within each cohort. Negative d indicates lower scores in the positive (impaired) group.

| Domain | MCI | ADpos |
|---|---|---|
| **BSHARP** | **(N: 308 vs 183)** | **(N: 129 vs 306)** |
| Articulatory Precision | -0.34 (0.09), <.001 | -0.58 (0.11), <.001 |
| Cognitive-Linguistic Domain | -0.22 (0.09), 0.02 | -0.28 (0.11), 0.009 |
| Executive Fluency & Planning | -0.29 (0.09), 0.002 | 0.26 (0.11), 0.02 |
| Global NST | -0.52 (0.09), <.001 | -0.40 (0.11), <.001 |
| Lexical-Semantic Domain | -0.57 (0.10), <.001 | -0.58 (0.11), <.001 |
| Morphosyntactic Complexity | -0.29 (0.09), 0.002 | -0.05 (0.10), 0.61 |
| Phonation & Laryngeal Control | 0.05 (0.09), 0.62 | -0.16 (0.11), 0.14 |
| Prosodic Modulation | -0.01 (0.09), 0.89 | 0.17 (0.11), 0.10 |
| **DHMS** | **(N: 196 vs 497)** | **(N: 195 vs 401)** |
| Articulatory Precision | 0.07 (0.08), 0.40 | 0.09 (0.09), 0.30 |
| Cognitive-Linguistic Domain | -0.21 (0.08), 0.01 | -0.19 (0.09), 0.03 |
| Executive Fluency & Planning | -0.57 (0.09), <.001 | -0.03 (0.09), 0.70 |
| Global NST | -0.40 (0.09), <.001 | -0.12 (0.09), 0.18 |
| Lexical-Semantic Domain | -0.32 (0.08), <.001 | -0.15 (0.09), 0.10 |
| Morphosyntactic Complexity | -0.08 (0.08), 0.35 | -0.02 (0.09), 0.78 |
| Phonation & Laryngeal Control | 0.13 (0.08), 0.13 | 0.07 (0.09), 0.42 |
| Prosodic Modulation | -0.33 (0.08), <.001 | -0.16 (0.09), 0.08 |
| **CCARE** | **(N: 209 vs 68)** | **(N: 40 vs 214)** |
| Articulatory Precision | -0.01 (0.14), 0.96 | 0.08 (0.17), 0.64 |
| Cognitive-Linguistic Domain | 0.07 (0.14), 0.62 | -0.21 (0.17), 0.23 |
| Executive Fluency & Planning | -0.24 (0.14), 0.08 | 0.01 (0.17), 0.95 |
| Global NST | -0.05 (0.14), 0.70 | -0.17 (0.17), 0.32 |
| Lexical-Semantic Domain | -0.32 (0.14), 0.02 | 0.06 (0.17), 0.75 |
| Morphosyntactic Complexity | -0.28 (0.14), 0.04 | -0.17 (0.17), 0.32 |
| Phonation & Laryngeal Control | 0.11 (0.14), 0.43 | 0.03 (0.17), 0.86 |
| Prosodic Modulation | 0.08 (0.14), 0.56 | -0.25 (0.17), 0.15 |

*p < .05, **p < .01, ***p < .001 (uncorrected).

Abbreviations: MCI, mild cognitive impairment; ADpos, Alzheimer's disease biomarker positive; NST, Neurocognitive Speech Taxonomy.

Table S6. Per-Cohort Effect Sizes for NST Tier-2 Category Scores by Clinical Outcome.

| Category | MCI | ADpos |
|---|---|---|
| **BSHARP** | **(N: 308 vs 183)** | **(N: 129 vs 306)** |
| Abstract Reasoning | -0.04 (0.09), 0.64 | 0.11 (0.11), 0.32 |
| Affective Expression | -0.02 (0.09), 0.85 | 0.06 (0.10), 0.57 |
| Attention Markers | -0.13 (0.09), 0.17 | -0.00 (0.10), 0.99 |
| Clause Structure | 0.32 (0.09), <.001 | 0.02 (0.10), 0.88 |
| Content Region Activation | 0.34 (0.09), <.001 | 0.16 (0.11), 0.13 |
| Disfluencies | 0.08 (0.09), 0.38 | 0.20 (0.11), 0.06 |
| Executive Markers | -0.11 (0.09), 0.25 | -0.00 (0.10), 0.97 |
| Fluency Disruption | 0.31 (0.09), 0.001 | 0.38 (0.11), <.001 |
| Formant Amplitude | 0.29 (0.09), 0.002 | 0.02 (0.10), 0.85 |
| Formant Energy | 0.28 (0.09), 0.003 | 0.03 (0.10), 0.75 |
| Formant Positioning | -0.34 (0.09), <.001 | 0.11 (0.11), 0.29 |
| Formant Precision | -0.32 (0.09), <.001 | 0.16 (0.11), 0.12 |
| Glottal Source Quality | -0.01 (0.09), 0.90 | -0.11 (0.11), 0.29 |
| Grammatical Composition | 0.39 (0.09), <.001 | 0.53 (0.11), <.001 |
| Intensity Dynamics | -0.18 (0.09), 0.05 | 0.03 (0.10), 0.79 |
| Intensity Level | -0.16 (0.09), 0.08 | 0.09 (0.11), 0.41 |
| Intensity Range | -0.22 (0.09), 0.02 | 0.16 (0.11), 0.12 |
| Lexical Composition | -0.31 (0.09), 0.001 | 0.09 (0.11), 0.41 |
| Memory Markers | -0.21 (0.09), 0.03 | -0.20 (0.11), 0.06 |
| Pause Behavior | 0.24 (0.09), 0.01 | 0.10 (0.11), 0.33 |
| Pause Patterns | 0.08 (0.09), 0.39 | -0.09 (0.11), 0.37 |
| Perceptual Processing | -0.22 (0.09), 0.02 | 0.01 (0.10), 0.96 |
| Pitch Dynamics | -0.15 (0.09), 0.10 | -0.14 (0.11), 0.19 |
| Pitch Range | -0.37 (0.09), <.001 | 0.15 (0.11), 0.16 |
| Pitch Stability | -0.22 (0.09), 0.02 | -0.16 (0.11), 0.12 |
| Repetitions | 0.05 (0.09), 0.59 | 0.53 (0.11), <.001 |
| Rhythmic Regularity | -0.37 (0.09), <.001 | 0.14 (0.11), 0.20 |
| Self-Interruptions | 0.14 (0.09), 0.13 | 0.32 (0.11), 0.003 |
| Semantic Fluency Output | -0.58 (0.10), <.001 | -0.41 (0.11), <.001 |
| Semantic Informativeness | -0.18 (0.09), 0.06 | -0.18 (0.11), 0.09 |
| Spectral Balance | -0.13 (0.09), 0.16 | 0.37 (0.11), <.001 |
| Spectral Dynamics | -0.31 (0.09), 0.001 | -0.16 (0.11), 0.13 |

| | | |
|---|---|---|
| Spectral Envelope | -0.07 (0.09), 0.43 | 0.07 (0.11), 0.52 |
| Speech Rate | 0.59 (0.10), <.001 | 0.76 (0.11), <.001 |
| Syntactic Span | -0.36 (0.09), <.001 | 0.14 (0.11), 0.18 |
| Temporal Density | -0.43 (0.09), <.001 | -0.14 (0.11), 0.18 |
| Vocabulary Breadth | -0.04 (0.09), 0.65 | 0.51 (0.11), <.001 |
| Vocal Stability | -0.23 (0.09), 0.01 | 0.12 (0.11), 0.25 |
| Voicing Segmentation | 0.02 (0.09), 0.82 | -0.19 (0.11), 0.06 |
| Word Complexity | -0.42 (0.09), <.001 | -0.06 (0.10), 0.59 |
| **DHMS** | **(N: 196 vs 497)** | **(N: 195 vs 401)** |
| Abstract Reasoning | -0.39 (0.08), <.001 | -0.09 (0.09), 0.30 |
| Affective Expression | -0.33 (0.08), <.001 | -0.11 (0.09), 0.20 |
| Attention Markers | -0.52 (0.09), <.001 | -0.15 (0.09), 0.09 |
| Clause Structure | -0.10 (0.08), 0.22 | -0.08 (0.09), 0.36 |
| Content Region Activation | 0.04 (0.08), 0.60 | 0.11 (0.09), 0.21 |
| Disfluencies | 0.05 (0.08), 0.52 | -0.07 (0.09), 0.45 |
| Executive Markers | -0.40 (0.09), <.001 | -0.15 (0.09), 0.10 |
| Fluency Disruption | -0.43 (0.09), <.001 | -0.04 (0.09), 0.64 |
| Formant Amplitude | -0.33 (0.08), <.001 | 0.00 (0.09), 0.96 |
| Formant Energy | -0.35 (0.08), <.001 | -0.01 (0.09), 0.87 |
| Formant Positioning | -0.18 (0.08), 0.04 | -0.13 (0.09), 0.15 |
| Formant Precision | -0.09 (0.08), 0.31 | -0.18 (0.09), 0.04 |
| Glottal Source Quality | -0.01 (0.08), 0.93 | -0.01 (0.09), 0.94 |
| Grammatical Composition | 0.06 (0.08), 0.45 | 0.04 (0.09), 0.63 |
| Intensity Dynamics | -0.10 (0.08), 0.22 | 0.02 (0.09), 0.84 |
| Intensity Level | 0.18 (0.08), 0.03 | 0.08 (0.09), 0.35 |
| Intensity Range | -0.22 (0.08), 0.01 | -0.09 (0.09), 0.33 |
| Lexical Composition | -0.30 (0.08), <.001 | -0.04 (0.09), 0.65 |
| Memory Markers | -0.55 (0.09), <.001 | -0.07 (0.09), 0.40 |
| Pause Behavior | 0.49 (0.09), <.001 | -0.06 (0.09), 0.48 |
| Pause Patterns | 0.51 (0.09), <.001 | -0.01 (0.09), 0.88 |
| Perceptual Processing | -0.64 (0.09), <.001 | -0.13 (0.09), 0.13 |
| Pitch Dynamics | -0.07 (0.08), 0.38 | -0.08 (0.09), 0.35 |
| Pitch Range | -0.08 (0.08), 0.35 | -0.07 (0.09), 0.40 |
| Pitch Stability | 0.19 (0.08), 0.02 | 0.09 (0.09), 0.30 |
| Repetitions | -0.51 (0.09), <.001 | -0.06 (0.09), 0.48 |
| Rhythmic Regularity | -0.28 (0.08), <.001 | -0.08 (0.09), 0.37 |

| | | |
|---|---|---|
| Self-Interruptions | -0.11 (0.08), 0.20 | 0.03 (0.09), 0.74 |
| Semantic Fluency Output | -0.19 (0.08), 0.03 | -0.15 (0.09), 0.09 |
| Semantic Informativeness | -0.52 (0.09), <.001 | -0.06 (0.09), 0.50 |
| Spectral Balance | 0.05 (0.08), 0.53 | 0.02 (0.09), 0.79 |
| Spectral Dynamics | 0.16 (0.08), 0.06 | 0.12 (0.09), 0.19 |
| Spectral Envelope | -0.12 (0.08), 0.17 | 0.15 (0.09), 0.09 |
| Speech Rate | -0.31 (0.08), <.001 | -0.03 (0.09), 0.76 |
| Syntactic Span | -0.40 (0.09), <.001 | 0.05 (0.09), 0.55 |
| Temporal Density | -0.52 (0.09), <.001 | -0.19 (0.09), 0.03 |
| Vocabulary Breadth | -0.63 (0.09), <.001 | -0.01 (0.09), 0.86 |
| Vocal Stability | -0.10 (0.08), 0.22 | -0.09 (0.09), 0.31 |
| Voicing Segmentation | -0.28 (0.08), <.001 | 0.07 (0.09), 0.43 |
| Word Complexity | -0.16 (0.08), 0.06 | -0.06 (0.09), 0.47 |
| **CCARE** | **(N: 209 vs 68)** | **(N: 40 vs 214)** |
| Abstract Reasoning | -0.10 (0.14), 0.49 | 0.04 (0.17), 0.80 |
| Affective Expression | -0.10 (0.14), 0.50 | -0.14 (0.17), 0.42 |
| Attention Markers | -0.25 (0.14), 0.07 | -0.19 (0.17), 0.28 |
| Clause Structure | 0.15 (0.14), 0.28 | 0.05 (0.17), 0.79 |
| Content Region Activation | -0.13 (0.14), 0.34 | -0.17 (0.17), 0.32 |
| Executive Markers | -0.22 (0.14), 0.11 | -0.20 (0.17), 0.24 |
| Formant Amplitude | 0.21 (0.14), 0.13 | -0.14 (0.17), 0.41 |
| Formant Positioning | -0.11 (0.14), 0.45 | -0.21 (0.17), 0.23 |
| Formant Precision | 0.03 (0.14), 0.83 | -0.17 (0.17), 0.32 |
| Glottal Source Quality | 0.05 (0.14), 0.73 | -0.16 (0.17), 0.35 |
| Grammatical Composition | 0.43 (0.14), 0.003 | 0.23 (0.17), 0.18 |
| Intensity Dynamics | -0.11 (0.14), 0.45 | 0.26 (0.17), 0.13 |
| Intensity Level | -0.13 (0.14), 0.35 | 0.24 (0.17), 0.17 |
| Intensity Range | 0.06 (0.14), 0.68 | -0.29 (0.17), 0.09 |
| Memory Markers | -0.28 (0.14), 0.05 | 0.06 (0.17), 0.71 |
| Pause Behavior | 0.05 (0.14), 0.74 | 0.16 (0.17), 0.36 |
| Pause Patterns | 0.02 (0.14), 0.88 | 0.17 (0.17), 0.33 |
| Perceptual Processing | -0.26 (0.14), 0.06 | -0.05 (0.17), 0.77 |
| Pitch Dynamics | -0.13 (0.14), 0.37 | -0.35 (0.17), 0.04 |
| Pitch Range | 0.18 (0.14), 0.20 | -0.20 (0.17), 0.25 |
| Repetitions | -0.43 (0.14), 0.002 | 0.07 (0.17), 0.69 |
| Rhythmic Regularity | -0.24 (0.14), 0.09 | 0.23 (0.17), 0.18 |

| | | |
|---|---|---|
| Semantic Informativeness | -0.32 (0.14), 0.02 | 0.06 (0.17), 0.72 |
| Spectral Balance | 0.01 (0.14), 0.93 | -0.10 (0.17), 0.58 |
| Spectral Dynamics | -0.04 (0.14), 0.76 | 0.30 (0.17), 0.08 |
| Spectral Envelope | -0.01 (0.14), 0.91 | 0.09 (0.17), 0.61 |
| Speech Rate | -0.28 (0.14), 0.05 | -0.13 (0.17), 0.44 |
| Syntactic Span | -0.46 (0.14), 0.001 | -0.21 (0.17), 0.22 |
| Vocabulary Breadth | -0.42 (0.14), 0.003 | 0.21 (0.17), 0.23 |
| Vocal Stability | 0.26 (0.14), 0.06 | -0.03 (0.17), 0.85 |
| Voicing Segmentation | -0.13 (0.14), 0.34 | 0.04 (0.17), 0.80 |
| Word Complexity | -0.40 (0.14), 0.004 | 0.12 (0.17), 0.50 |

Values are Cohen's d comparing outcome-positive vs outcome-negative groups for each tier-2 NST category within each cohort.

Tier-2 categories are finer-grained groupings within each tier-1 domain (e.g., Pitch Stability within Prosodic Modulation).

Abbreviations: MCI, mild cognitive impairment; ADpos, Alzheimer's disease biomarker positive; NST, Neurocognitive Speech Taxonomy.

Table S7. Racial and Educational Disparities in MoCA and NST Domain Scores.

| Measure | BSHARP Racial d | DHMS Racial d | BSHARP Education d | DHMS Education d |
|---|---|---|---|---|
| **MoCA** | 0.373 | 0.722 | 1.692 | 0.675 |
| **Articulatory Precision** | -0.210 | 0.082 | -0.085 | 0.306 |
| **Cognitive-Linguistic Domain** | 0.030 | 0.065 | 0.091 | 0.106 |
| **Executive Fluency and Planning** | 0.305 | 0.328 | 0.285 | 0.618 |
| **Lexical-Semantic Domain** | 0.129 | 0.205 | 0.634 | 0.249 |
| **Morphosyntactic Complexity** | 0.255 | 0.176 | 0.209 | 0.217 |
| **Phonation and Laryngeal Control** | -0.120 | -0.134 | 0.206 | 0.138 |
| **Prosodic Modulation** | 0.255 | 0.153 | -0.149 | 0.709 |

Cohen's d for racial disparity: White minus non-white group mean difference, standardized by pooled SD. Positive d indicates higher scores in the White group. Cohen's d for education disparity: high education (≥ median years) minus low education (< median years). Positive d indicates higher scores in the higher-education group.

MoCA shows substantially larger racial (d = 0.37–0.72) and educational (d = 0.68–1.69) disparities than most NST domain scores, suggesting speech-based measures may be less susceptible to sociodemographic bias.

CCARE excluded from racial analysis (100% non-white).

Abbreviations: d, Cohen's d; MoCA, Montreal Cognitive Assessment; NST, Neurocognitive Speech Taxonomy.
